# Adherence of SARS-CoV-2 delta variant to surgical mask and N95 respirators

**DOI:** 10.1101/2022.01.05.22268808

**Authors:** Ana C. Lorenzo-Leal, Selvarani Vimalanathan, Horacio Bach

## Abstract

The use of facial protection, including masks and respirators, has been adopted globally due to the COVID-19 pandemic. These products have been demonstrated to be effective in reducing the transmission of the virus. To determine whether or not the virus adheres to masks and respirators, we dissected four respirators and one surgical mask into layers. These individual layers were contaminated with the SARS-CoV-2 delta variant, and its release by vortexing was performed. Samples were used to infect Vero cells, and a plaque assay was used to determine to evaluate the adherence of the virus. Results showed that a cumulative log reduction of the layers reduced the load of the virus six-folds. Our study confirms the effectiveness of facial protection in reducing the transmission and or infection of the virus.

## 1. Introduction

The pandemic of the novel coronavirus SARS-CoV-2, the causative agent of COVID-19, was initially identified in Wuhan, China, and has spread worldwide. According to the World Health Organization (WHO), as of November 20^th^, 2021, there have been 5,127,696 deaths and 255,324,963 confirmed cases of COVID-19 [1].

COVID-19 disease seems to be spreading through airborne transmission via aerosol, and as it is known that respiratory infections mainly occur when breathing, speaking, sneezing, coughing, or talking occurs. These transmissions are facilitated by droplets (> 5 to 10 µm) and/or aerosols (≤ 5 µm) containing the virus SARS-CoV-2 (ref). Aerosol’s accumulation, also known as the “gas cloud,” could remain infectious for hours in indoor spaces. Thus, the implementation of masks or respirators has a tremendous effect on reducing aerosol transmission [2–5]. This physical protection is in use and has been adopted globally.

There are different types of masks or respirators with various levels of protection, which can be reusable or disposable. Generally, masks do not fit as tightly as respirators. For example, N95 respirators are classified by the U.S. National Institute for Occupational Safety and Health (NIOSH) as air filtration, filtering at least 95% of airborne particles. N95 respirators and surgical masks are examples of the most common disposable masks, while reusable are mainly industrial masks with half or full facepiece respirators [5].

According to the Centers for Disease Control and Prevention (CDC) and the NIOSH, surgical masks are approved by the Food and Drug Administration (FDA) and intended to use as protection to the wearer against multiple hazardous environments, such as those created by sprays, dangerous fluids, splashes, and droplets. However, masks would not always confer respiratory protection because of their lower reliable level of filtration against smaller particles that could be inhaled, such as airborne particles.

Surgical masks contain mainly three-layer structures with different functions. For example, the outer layer is repelling water, the middle layer functions as a trap of particles and prevents the penetration of microorganisms, and the inner layer absorbs moisture [2,6].

On the other hand, N95 respirators are defined by NIOSH as a filtration system that retains at least 95% of different airborne sizes particles (from aerosols to large droplets), reducing the exposure to the user [7]. Respirators may contain extra layers compared to surgical masks. For example, N95 respirators have between three to five layers, comprising an inner and outer layer, a filter layer, and in some cases, a support layer [6].

The penetration or filtration mechanism of aerosols containing SARS-CoV-2 in masks has been studied and reported in different studies [2,3,6,8,9]. Moreover, contamination with SARS-CoV-2 on the masks’ outer surface has also been reported [10]. Still, a detailed investigation addressing the question of whether or not the virus remains adhered to the layers has not been reported. Then, this study aimed to evaluate the adherence of the SARS-CoV-2 delta variant on different layers of a surgical mask and N95 respirators.

## 2. Methods

### 2.1. Virus and cell lines

SARS-CoV-2 delta variant (B.1.617.2) was provided by the British Columbian Centre for Disease Control Public Health Laboratory, Vancouver, Canada. Vero E6 cells (ATCC CCL-81) were used to replicate the virus and infections. Cells were grown in MEM (Invitrogen) supplemented with 5% fetal calf serum (Invitrogen), pyruvate, and non-essential amino acids. All the incubations were performed in an incubator at 37°C supplemented with 5% CO_2_. All the work performed in this study was conducted in level 3 at the Facility for Infectious Disease and Epidemic Research (University of British Columbia, Vancouver, BC, Canada).

Viral stocks were prepared by infecting 80% confluent Vero E6 cells with 1 mL of the virus at a 5×10^5^ PFU/mL concentration. After 72 h, the supernatant was collected and cleared by centrifugation at 3200 × *g* for 10 min. The supernatant was aliquoted and kept at −80°C until further use. The titer of the virus was determined to be 5.9×10^5^ pfu/mL by plaque assay.

A plaque assay was performed overnight by dispensing 1.5×10^5^ Vero cells into 12-well plates. The next day, the 80-100% confluency was verified, and cells were infected with the virus for one h. Then, the virus was washed away, and the monolayer was overlayed with 1.5 mL of 2 x MEM supplemented with 2% cellulose (Sigma) (v/v 1:1)

### 2.2. Mask or respirators preparation

Five different masks or respirators (Table 1) were used in this study. Pieces of 1 cm^2^ were cut, separated into individual layers, decontaminated by submerging the pieces into 70% ethanol for 10 min, and dry overnight at 37 °C inside a decontaminated Biosafety Containment Level 2. The decontaminated layers were stored in empty sterile 1.5 mL tubes until further use.

**Table 1.** Characteristics of the masks and respirators used in this study

| Brand | Layer | Material | Reference |
| --- | --- | --- | --- |
| 1 North N95 7130 (Lot No. 9E001)<br>Closed-cell vinyl foam | Outer<br>Filter<br>Support<br>Inner | Polypropylene<br>Polypropylene composite<br>Polyester<br>Polyester | [11] |
| **2** North N95 5130 (Lot No. 9G004)<br>Open-cell foam seal | Outer<br>Filter<br>Inner | Polyester<br>Polypropylene composite<br>Polyester | [11] |
| **Three** 3-Ply surgical mask | Outer<br>Filter<br>Inner | Fluid resistant<br>Polypropylene<br>Absorbent | [12] |
| **4** 3M 8210 N95 (Lot No. A20047)<br>Standard respirator | Outer<br>Filter<br>Inner | Polyester<br>Polypropylene<br>Polyester | [13] |
| **5** 3M 1860 N95 (Lot No. B16047)<br>Surgical respirator | Outer<br>Filter<br>Inner | Polyester<br>Polypropylene<br>Polyester | [13] |

### 2.3. Contamination of layers

The layers were exposed to 2 × 10^3^ viruses (in 10 μL MEM). After 10 min (time to dry the drop), the layers were submerged in 300 µL of OptiMEM (Invitrogen) in a 1.5 mL screw-top tube and vortexed vigorously for 1 min every. The samples were subjected to 10-fold dilutions using OptiMEM in 1.5 mL tubes.

### 2.4. Plaque assay

5.0 × 10^5^ Vero E6 cells in complete MEM were seeded into 12-well cell culture plates (Thermo Scientific) and incubated overnight at 37 °C supplemented with 5% CO_2_. Cells were then checked for confluency, and the medium was removed from the wells. The serial dilutions (300 μL) were added to the corresponded wells, and the plates were incubated as explained above. The plates were placed back in the incubator for an hour to allow infection. The plates were gently rocked every 15 min. Then, the virus and the medium were removed from the wells, and an overlay of 2% cellulose and 2x complete MEM (as detailed above) was gently added to each well. Plates were incubated for 72 h, fixed for 30 min with 4% *p*-formaldehyde (PFA, J.T. Baker) in PBS. After removing the PFA, the cells were stained with 250 μL of 1% crystal violet (Sigma), dissolved in 20% methanol (Fisher) for 3,0 min, washed twice with water, dried, and the number of plaques was counted and recorded.

## 3. Results and discussion

The adherence of SARS-CoV-2 to different types of buccal protection showed that all of the layers could retain the virus at different extents. In addition, adherent viruses were not released upon vortexing the samples, suggesting that they are tightly bound to the layers once they are deposited and dried on their surface.

Analysis of the results showed that an accumulation of the log reductions of the layers ranged between 6.15-8.45 (Figure 2). In addition, respirator **2** showed superior retention of the virus with an increase of approximately 1-2 log reduction (Figure 2B) compared to the rest of the tested products.

**Figure 1.**
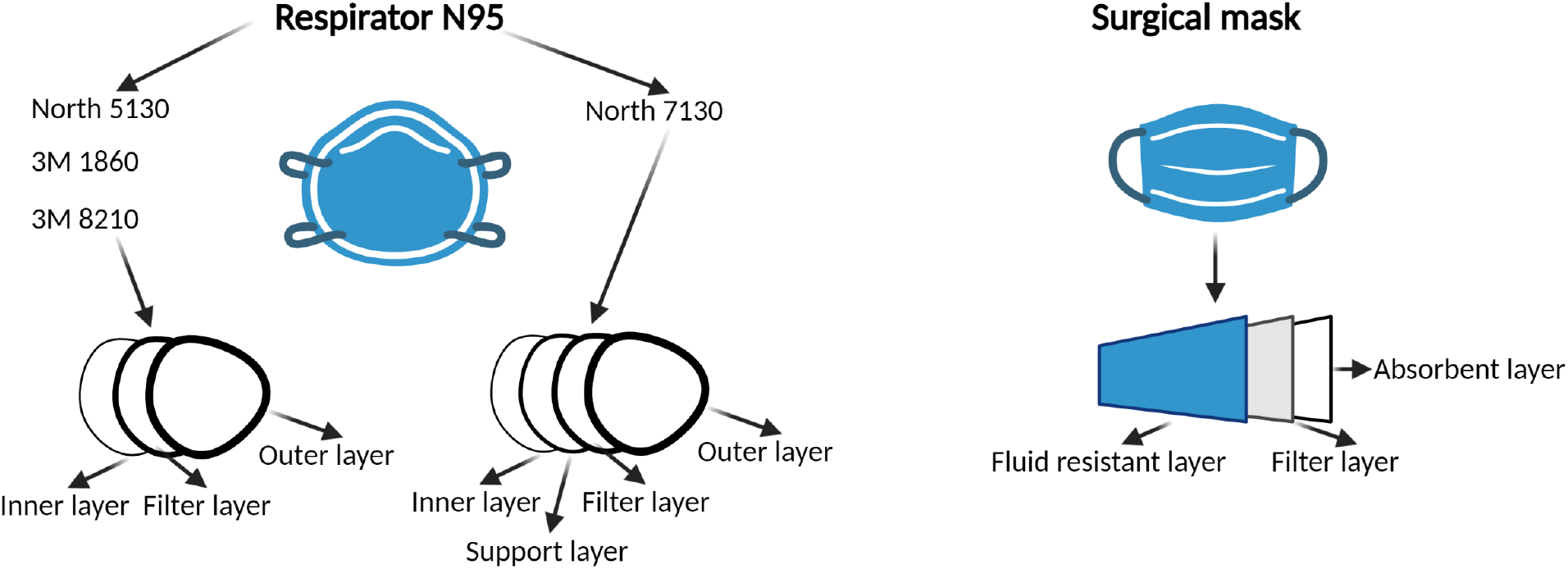
Mask or respirators layers. Description of the mask and respirator layers used in this study. Created by Biorender.com

**Figure 2.**
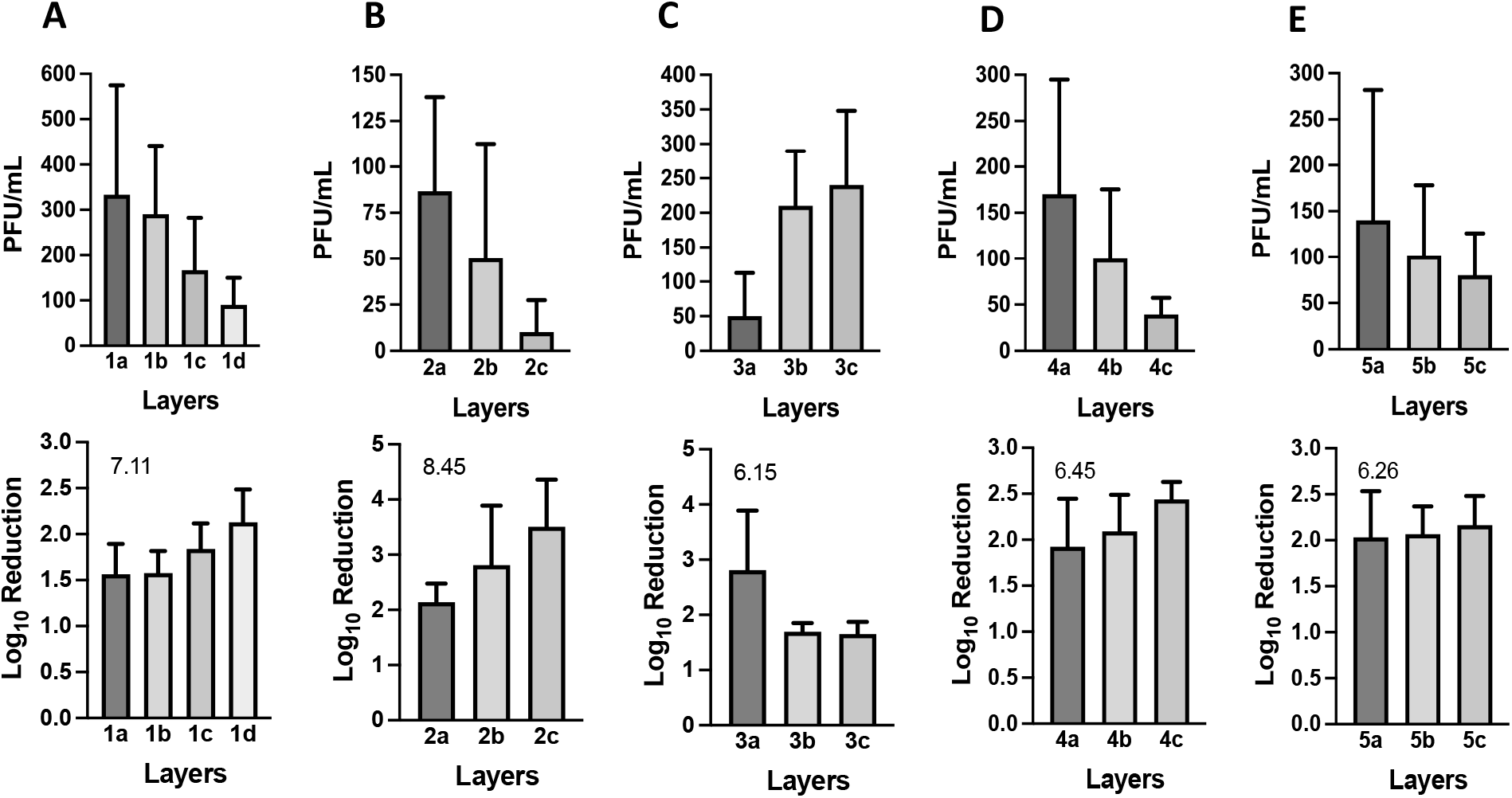
Viral titers of SARS-CoV-2 deposited onto layers. The virus was deposited on the layers, and after drying, non-adherent viruses were released from the layers by vortexing and according to the Method section. (A) Respirator 7130, (B) Respirator 5130, (C) Surgical masks, (D) Respirator 8210, and (E) Respirator 1860. Shown is the mean of at least three independent experiments ± SD. The number located in the lower panel is the cumulative log reduction of the layers. The arrangement of the layers is from the outer layer (a) to the inner layer (c), except for **one** that has four layers (d).

People use medical masks as protection from sprays and splashes of fluids by sick people. Although extended usage or reuse of masks has become common, especially during the present pandemics, this could lead to an infection of the person wearing the mask due to the possible presence of pathogens in different layers of the masks [10].

Masks materials (Table 1) are mainly nonwoven fabric made by a mass of fibers (such as polypropylene, polyester, and polyurethane) bonded by different means such as mechanical, heat or chemical. The efficiency of particle filtration (permeability), bacterial or virus filtration, differential pressure, flammability, flexibility, and resistance to fluids are some of the performance characteristics for materials used in masks [2,3,6]. Regarding permeability, when resistance to fluid material is used, aerosols are likely to travel around the mask following contours seeking low resistance areas or leeks, rather than penetration of the filter material [3].

Between the different layers in a mask or respirator, the most important one is the filter layer, which is mainly produced through a melt-blown process, where a polymer (such as polyethylene) is exposed to blows of high-velocity air, forming a web shape of filaments. This filter layer (commonly used in N95 respirators) has an electrostatic charge; therefore, the polymer filtration capability for small particles increases because of electrostatic adsorption [2,14]. In our study, the respirators contained at least one layer of polypropylene, suggesting that an electrostatic layer increased the retention of the virus.

A model of viral retention based on the electrostatic charges on the surface of the virus and the modified polypropylene (electret) might explain one of the reasons for the log reduction of the virus. In the case of the virus, the Spike protein surrounds the viral particle. Assuming that other components of the virus are not contributing to the surface charge of the virus (lipids and nucleocapsid are part of the core of the virus [15], we can attribute the net surface charge of the virus to the Spike protein. This protein looks to have a particular disorder [16] probably because of its interaction with the membrane proteins [17]. Thus, the electrostatic interaction of the virus and the electret is based on the charge of the Spike protein [18]. One of the main variables regarding the isoelectric point of the Spike protein is the large number of ionizable amino acids that contribute to the net charge (e.g., C, D, E, H, K, R, and Y) [15]. Then, the pH will play an important role in the fluctuation of the surface charge at which the virus.

The isoelectric point of the Spike protein is 6.2 [19]. Although this value is lower than the physiological pH (7.0), most Spike protein is negatively charged [15], except the receptor-binding domain. This receptor is located in the apical portion of the spike protein, and it has been calculated to remain positively charged through a range of pH values [20]. Interestingly, the positive charge of the receptor-binding domain has increased in the SARS-CoV-2 variants [20] because of the mutation observed in this region.

On the other hand, studies reported negative surface charges on the polypropylene electrets, varying between −200 to −1350 (V) [21,22]. Thus, a strong electrostatic attraction occurs between the receptor-binding domain of the Spike protein and the polypropylene electret, contributing to the retention of the viruses on the layers.

## 4. Conclusions

In this study, we report the adherence of the SARS-CoV-2 delta variant to different buccal protection (mask and respirators). We hypothesize that a strong electrostatic binding occurs, and as a result, the cumulative viral log reduction is over 6. Our results also showed that masks and respirators efficiently reduce infections either from the infected (wearer) or the recipient person.

## Data Availability

All data produced in the present work are contained in the manuscript

## Acknowledgement

We thank Dr. Mel Krajden from the British Columbian Centre for Disease Control Public Health Laboratory, Vancouver, Canada for the access to the SARS-CoV-2 delta variant.

## Author Contributions

H.B. conceptualized, reviewed, edited the manuscript, and procured financing acquisition. A.L. investigated, wrote, reviewed, and edited the manuscript. S.V. investigated and reviewed the manuscript. All authors have read and agreed to the published version of the manuscript.

## Funding

This work was supported by the Canadian Institutes of Health Research (CIHR) grant Canadian 2019 Novel Coronavirus (COVID-19) Rapid Research Funding Opportunity Therapeutics to H.B (No. OV3-170627).

## Conflicts of Interest

The authors have nothing to disclose.

## Notes

### Competing Interest Statement

The authors have declared no competing interest.

